# Effects of Age and Knee Osteoarthritis on the Modular Control of Walking: A Pilot Study

**DOI:** 10.1101/2020.05.22.20110536

**Authors:** Sarah A. Roelker, Rebekah R. Koehn, Elena J. Caruthers, Laura C. Schmitt, Ajit M.W. Chaudhari, Robert A. Siston

**Affiliations:** Department of Kinesiology, University of Massachusetts Amherst, Amherst, MA, USA; Department of Mechanical and Aerospace Engineering, The Ohio State University, Columbus, OH, USA; Department of Engineering, Otterbein University, Westerville, OH, USA; School of Health and Rehabilitation Sciences, The Ohio State University, Columbus, OH, USA; Sports Medicine Research Institute, The Ohio State University Wexner Medical Center, Columbus, OH, USA; Division of Physical Therapy, School of Health and Rehabilitation Sciences, The Ohio State University, Columbus, OH, USA; Department of Biomedical Engineering, The Ohio State University, Columbus, OH, USA; Department of Orthopaedics, The Ohio State University, Columbus, OH, USA

**Keywords:** muscle synergy, motor control, co-activation, electromyography, gait

## Abstract

Older adults and individuals with knee osteoarthritis (KOA) often exhibit reduced locomotor function and altered muscle activity. Identifying age- and KOA-related changes to the modular control of gait may provide insight into the neurological mechanisms underlying reduced walking performance in these populations. The purpose of this pilot study was to determine if the modular control of walking differs between younger and older adults without KOA and adults with end-stage KOA. Kinematic, kinetic, and electromyography data were collected from ten younger (23.5 ± 3.1 years) and ten older (63.5 ± 3.4 years) adults without KOA and ten adults with KOA (64.0 ± 4.0 years) walking at their self-selected speed. Separate non-negative matrix factorizations of 500 bootstrapped samples determined the number of modules required to reconstruct each participant’s electromyography. The number of modules required in the younger adults (3.2 ± 0.4) was greater than in the individuals with KOA (2.3 ± 0.7; *p* = 0.002), though neither cohorts’ required number of modules differed significantly from the unimpaired older adults (2.7 ± 0.5; *p* ≥ 0.113). A significant association between module number and walking speed was observed (*r* = 0.532; *p* = 0.003) and individuals with KOA walked significantly slower (0.095 ± 0.21 m/s) than younger adults (1.24 ± 0.15 m/s; *p* = 0.005). Individuals with KOA also exhibited altered module activation patterns and composition (which muscles are associated with each module) compared to unimpaired adults. These findings suggest aging alone may not significantly alter modular control; however, the combined effects of knee osteoarthritis and aging may together impair the modular control of gait.

## INTRODUCTION

Functional limitations, including difficulty walking, climbing stairs, or crouching, in community dwelling older adults aged have been shown to be predictive of future disability (1) as well as falls, pain, and medical expenses (2). Age-related change in muscle function has been identified as the most important physiological change leading to functional limitations (3). Impaired neuromuscular activity has been identified as a cause of this altered muscle function (4,5). Furthermore, age-related differences in muscle activation patterns have been linked to altered gait kinematics and kinetics in older adults with no history of musculoskeletal injury or joint disorders (6–9). Older adults exhibit highly repeatable electromyography (EMG) signals, which suggests an inability to adapt their motor control to perturbations and reflects a loss of neural plasticity (6,9). This age-related decrease in control complexity and adaptability of the nervous system may indicate that changes to the organization of the neural mechanisms underlying motor control are responsible for impaired functional performance in older adults.

Furthermore, it is important to differentiate between neuromuscular changes associated with normal aging and those associated with age-related disorders. For example, symptomatic knee osteoarthritis (KOA) is a leading cause of disability in older adults (10); yet, not all older adults develop KOA. Individuals with KOA have difficulty completing activities of daily living (11) and walk with altered kinematics and kinetics compared to unimpaired age-matched individuals, including decreased peak knee flexion angle and peak knee flexion and extension moments (12). KOA-related changes in joint mechanics have been suggested to be related to altered muscle activation patterns in the presence of KOA (13).

Several studies have identified altered muscle activation patterns in individuals with KOA, which are suggestive of a disease-related loss of motor control complexity that may contribute to the observed changes in joint mechanics. For example, individuals with KOA exhibit increased co-contraction of the vastus medialis and medial gastrocnemius compared to unimpaired controls (14). In addition, principal component analyses of vasti, hamstring, and gastrocnemius activation patterns from asymptomatic adults and individuals with mild to moderate (Kellgren-Lawrence (KL) grades I-III (15)) (16,17) and severe (KL IV) KOA (17,18) revealed that a similar principal activation pattern captured the general spatiotemporal activation pattern of each muscle group across subject groups. However, subtle differences in the magnitude and shape of the activation patterns were capable of distinguishing between asymptomatic and KOA groups (16,17,19) and between KOA groups with different levels of structural severity (17,19). These differences between KOA and asymptomatic individuals’ activation patterns may indicate changes in motor control strategy with increasing disease severity.

Current motor control theory suggests the underlying mechanisms controlling muscle activity during gait can be decomposed into a few “primitive signals” (20–22) that represent basic central features of the motor programs (20). Muscles are grouped into modules (sometimes referred to as synergies) based on the similarity of their activation patterns to the temporal pattern of the primitive signals. A greater number of modules required to represent the original muscle activation patterns suggests a more complex neuromuscular control strategy (23). Weighting factors are assigned to the modules to quantify the strength of each muscle’s representation within a given module. Modular organization (muscle weightings and temporal patterns) has been shown to be consistent across walking speeds in healthy adults (22,23), which indicates these modules are a robust representation of the underlying neuromuscular control of gait (24).

Although modular control of gait has been characterized in healthy younger adults (age range: 26 to 42 years) (22,25) and healthy older adults (63.1 ± 9.1 years) (23,24,26), differences in the factorization method and the number of muscles included in the factorization used to identify modules in younger and older adults may limit accurate comparison of the modular control of gait between age-groups. For example, by using experimental EMG from 16 muscles, five modules were previously identified to control gait in younger adults (22,25). In contrast, four modules were identified in healthy older adults from the EMG of 8 muscles (23) while forward dynamic simulations of gait identified a fifth (24) and sixth (26) module that describe the three-dimensional modular control of gait. While a qualitative comparison of the modules identified in these individual studies suggest the modular control of gait in older and younger adults is characterized by similar temporal activation patterns and muscle weighting modules, only one study by Monaco et al. (27) has directly compared the modular control of gait between age-groups. Although Monaco et al. found no significant differences in module muscle weightings and temporal patterns between younger and older adults during gait, the authors prescribed the number of modules extracted to be equal to 5 for all participants prior to performing their factor analysis, which may have hindered their ability to identify differences in modular control between age-groups.

Alternatively, other studies of age-related differences in neuromuscular control suggest older adults utilize altered modular control strategies. Investigations of age-related differences in muscle synergies controlling postural responses during step preparation demonstrated diminished control of gait initiation in older adults compared to younger adults (28,29), which suggests that aging may alter modular control of locomotor tasks. In addition, analyses of upper limb muscle modules during a pointing task found reduced modular control complexity (fewer modules) in older adults compared to younger adults (30). Modular control analyses in populations with neuromuscular impairments, such as stroke and cerebral palsy, reveal reduced motor control complexity during both upper extremity tasks (31–33) and gait (23,34), which suggests populations exhibiting altered neuromuscular control in upper limb movements may also exhibit altered control of locomotion.

One recent study investigated the modular control of patients who had undergone a total knee arthroplasty (TKA), the end-stage treatment for KOA (35). Greater modular control complexity was observed in high-functioning compared to low-functioning TKA patients, although both TKA groups exhibited reduced complexity compared to unimpaired controls (35). However, whether modular control was altered due to the disease prior to surgery or was altered as a result of surgery is unknown. It is possible that the observed differences between KOA and asymptomatic individuals’ muscle activation patterns (16,17,19) are due to changes in the underlying motor control strategy.

Together, the altered muscle activity observed in older adults compared to younger adults and in individuals with KOA compared to unimpaired older adults suggests that modular control may differ between age groups as well as between healthy and impaired older adults. Identifying age- and pathology-related changes to the modular control of gait may provide insight into the neurological mechanisms underlying reduced locomotor performance. Therefore, the purpose of this pilot study was to determine if the modular control of gait differs between healthy younger and older adults and between healthy older adults and individuals with end-stage KOA. We hypothesized that there would be no differences in locomotor complexity, quantified by the number of modules required to reconstruct the experimental activation patterns, between younger and older adults without KOA, but that the modular organization of the older adults would be different than the younger adults. In addition, we hypothesized that the muscle activation patterns of individuals with end-stage KOA would reduce to fewer modules than that of the younger and older adults without KOA, representing a reduction in neuromuscular control complexity.

## MATERIALS AND METHODS

### Data Collection

Data from ten younger adults (5 female; 23.5 ± 3.1 years; 1.8 ± 0.1 m; 71.2 ± 10.0 kg), ten unimpaired older adults (5 female; 63.5 ± 3.4 years; 1.7 ± 0.1 m; 71.5 ± 13.5 kg), and ten individuals with primarily medial compartment KOA who were scheduled for a total knee arthroplasty within the following 8 weeks (7 female; 64.0 ± 4.0 years; 1.7 ± 0.1 m; 89.7 ± 7.5 kg) were included in this study as a secondary analysis of previously collected data. All participants provided written informed consent prior to participating in the data collection. All study procedures were approved by The Ohio State University Institutional Review Board. All participants performed at least five over-ground walking trials at their self-selected speeds. Self-selected speed was calculated as the distance between heel-markers during consecutive ipsilateral heel-strikes divided by the time between heel-strikes. Skin-mounted reflective markers were applied to the upper and lower extremities according to the modified Full-Body Point-Cluster Technique (PCT) to measure full body kinematics (36,37). The three-dimensional trajectories of the reflective markers were collected at 150 Hz using ten Vicon MX-F40 cameras (Vicon, Los Angeles, CA). An embedded-force-plate walkway formed by 6 force platforms (Bertec, Columbus, OH) collected ground reaction forces (GRFs) at 1500 Hz. Surface electromyography (EMG) (Telemyo DTS System, Noraxon, Scottsdale, AZ) was collected at 1500 Hz from 8 muscles on each participant’s dominant (younger and older adults) or involved (KOA) limb to measure muscle activation patterns from the rectus femoris, vastus lateralis, vastus medialis, biceps femoris, medial hamstrings, lateral gastrocnemius, medial gastrocnemius, and soleus. The skin over each muscle was prepared by shaving hair from the skin and cleansing the area with alcohol. Ag/AgCl dual-electrodes (Vermed, Buffalo, NY, Peripheral Nerve Stimulation Dual Element Electrodes, rectangular, 1.625 in x 3.25 in, 0.42 in sensor diameter, 1.625 in inter-electrode distance) were placed over the belly of each muscle. To determine the dominant limb of the healthy younger and older adults, each participant was asked which leg he or she would use to kick a ball (38).

### KOA-Specific Data Collection

The individuals with KOA included in this study are a subset of participants from a larger study investigating gait biomechanics before and after total knee arthroplasty (39,40). For each individual with KOA, two fellowship-trained, musculoskeletal radiologists graded the patient’s tibiofemoral radiographic severity using the KL grading system (15) by consensus. In addition to the walking trials, the individuals with KOA performed three clinical performance-based assessments: the six-minute walk test (6MW) (41), the timed stair climbing test (SCT) (42), and the timed-up and go test (TUG) (43). Finally, self-reported function was evaluated using four subscales of the Knee Injury and Osteoarthritis Outcome Score (KOOS) (44): pain, symptoms, activities of daily living (ADL), and knee-related quality of life (QOL). Each subscale is evaluated independently with scores between 0 and 100, with higher scores indicating better self-perceived function.

### Identification of Muscle Modules

Nonnegative Matrix Factorization (NMF) (45) was chosen to identify modules because it produces additive components that can be interpreted physiologically (46) given that neuron firing rates are never negative and synaptic strengths do not change sign (45). For each participant, EMG from a minimum of 5 gait cycles (mean: 11.3 ± 4.8 cycles; range: 5 – 23 cycles) were included in the analysis. The EMG data were demeaned, passed through a 30-300 Hz sixth order Butterworth band-pass filter, and full wave rectified. The data were then passed through a sixth order Butterworth low-pass filter at 6 Hz to create linear envelopes. Each muscle’s processed EMG data were normalized to the maximum EMG activation value from all walking trials. For each gait cycle, the normalized EMG was time normalized to 201 time points, accounting for every 0.5% of the gait cycle. Then, the normalized EMG from all gait cycles, *g*, were concatenated into an *m x t* matrix (EMG_o_), where *m* indicates the number of muscles (8) and *t* indicates the total number of time points (201 x *g*). Each muscle’s concatenated EMG in EMG_o_ was normalized to unit variance. An NMF algorithm defined by Lee and Seung (1999) was applied to the *m x t* matrix. NMF defines the modular organization by populating an *m x n* matrix representing the relative weighting of each muscle within each module, *n*, (Weighting Matrix) and an *n x t* matrix reflecting the temporal activation of the module across the gait cycle (Pattern Matrix). NMF assumes that muscles may belong to more than one module and that the muscle weightings on each module are fixed throughout the gait cycle. The two matrices were multiplied to produce an *m x t* matrix of the reconstructed activation patterns (EMG_r_). A multiplicative update algorithm using 50 replicates and a maximum of 10000 iterations adjusted the muscle weightings and activation profiles to minimize the sum of squared errors between EMG_o_ and EMG_r_.

For each participant in each population, separate NMF analyses extracted *n* modules from EMG_o_, where *n* ranged from 1 to 8 modules. For each extraction, the percent variability accounted for (VAF) by EMG_r_ was calculated as a measure of the agreement between the original and reconstructed activation patterns. The VAF for each muscle, *m*, (mVAF) was calculated as:

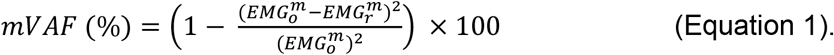

The total VAF (tVAF) was calculated as:

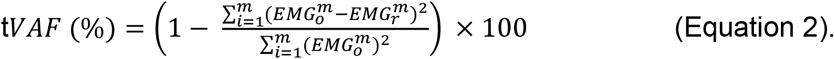

The 95% confidence interval (CI) of the VAF was determined using a bootstrapping procedure (47). For each number of modules extracted (1 to 8), the EMG_o_ matrix was resampled 500 times with replacement. For each sample, the muscle mVAFs and the tVAF were calculated and then the 95% CIs of the mVAFs and tVAF were constructed from the bootstrapped VAF values. The number of modules required to adequately reconstruct EMG_o_ was chosen as the minimum number of modules extracted for which the lower bound on the 95% CI for the tVAF was greater than or equal to 90% and the minimum mVAF was greater than 75% or adding another module did not increase the minimum mVAF by more than 5%.

### Module Analsyis

Module composition (which muscles are primarily associated with each module) was characterized by the Weighting Matrices from the bootstrapped samples. Since NMF outputs the modules in a random order for each bootstrap sample, the *n* columns of each Weighting Matrix associated with each *n* module must be sorted to ensure consistent module characterization across samples (e.g., the first column/module is always associated with high quadriceps activity, the second column/module is always associated with high plantarflexor activity, etc.). To sort the modules, *k*-means clustering was used to classify the *n* column of each sample’s Weighting Matrix associated with each module using the Weighting Matrix extracted by the NMF from EMG_o_ as the seed. On a sample-by-sample basis, each module’s muscle weightings were normalized to the maximum muscle weighting in the module, which resulted in a muscle weight of 1 for the muscle which had the greatest weighting in the module. Similarly, each participant’s principal patterns were normalized to the peak value so that the magnitude of each pattern ranged from 0 to 1. Then, for each module, the muscle weightings were averaged over the 500 bootstrapped samples so that 95% CIs could be identified for the muscle weightings.

Within each population, common modules between participants were identified using Pearson’s correlation coefficients. Modules from two participants were considered common if the module weightings were correlated with *r* ≥ 0.834, which corresponds to the critical *r*^2^ for 8 muscles at *p* = 0.01 (48). For each module that was common between participants in a population, the module muscle weightings were averaged across all participants with that common module. Between population correlations of average muscle weightings for common within population modules were determined to quantify similarity in module composition across populations. Module composition was also compared between populations based on the number of significantly active muscles per module (W_musc_) and the sum of the contributions of the significantly active muscles in a module (W_sum_; i.e., the sum of the bar heights of the muscle weightings) (49). Both W_musc_ and W_sum_ quantify the muscle co-activity of the module. A muscle was considered significantly active if the 95% CI for the muscle weighting did not include zero. The activation timing profiles of common modules were compared using the area under the activation curve (AUC). For each participant, the average activation timing profile from the *g* gait cycles included in the analysis was determined for each common module. Then, the AUC was calculated for six gait phases: weight acceptance (0-15% gait cycle), early midstance (15-30% gait cycle), late midstance (30-50% gait cycle), terminal stance (50-65% gait cycle), early swing (65-80% gait cycle), and late swing (80-100% gait cycle).

### Joint Kinematic and Kinetic Analysis

Sagittal plane joint angle and internal joint moment profiles for the hip, knee, and ankle were determined for each participant from a representative gait cycle and then averaged within each population. Joint moments were normalized by body weight, height, and self-selected walking speed. At the hip, peak angles were calculated for early stance hip flexion (H1), late stance hip extension (H2), and swing phase hip flexion (H3) and peak moments were calculated for early stance hip extension (HM1), late stance hip flexion (HM2), and swing phase hip extension (HM3). At the knee, peak angles were calculated for early stance knee flexion (K1), midstance knee extension (K2), and swing phase knee flexion (K3) and peak moments were calculated for weight acceptance knee flexion (KM1), early stance knee extension (KM2), midstance knee flexion (KM3), late stance knee extension (KM4), and swing phase knee flexion (KM5). At the ankle, peak angles were calculated for early stance plantarflexion (A1), late stance dorsiflexion (A2), push-off plantarflexion (A3), and swing phase dorsiflexion (A4); and peak moments were calculated for early stance dorsiflexion (AM1) and late stance plantarflexion (AM2). Peak measures were averaged across participants within each population.

### Statistics

Individual one-way Analysis of Variance (ANOVA) tests were used to assess the effect of group on age, height, mass, body mass index (BMI), self-selected walking speed, stride length normalized to leg length, the number of modules required to adequately reconstruct the EMG, and peak joint angles and moments. Individual one-way ANOVAs assessed the effect of group on W_musc_, W_sum_, AUC for common modules between populations. Individual one-way ANOVAs assessed the effect of group on peak joint kinematics and kinetics. When appropriate, Bonferroni post-hoc analyses were used to test for pairwise differences. The effect of the absence of a common module typically found in a healthy young adult (due to merging with another module) on peak joint kinematics and kinetics was assessed by two-sample t-tests between those participants with and without an independent common module, regardless of population. Participants who did not have a common module because the control of the primary muscles in the module was separated into two modules were excluded from this particular analysis. For the set of all participants and each population individually, Kendall rank correlations (Kendall’s *τ*_*b*_) assessed the association between the number of required modules and self-selected walking speed. For the KOA group, Kendall rank correlations were also used to assess the association between the number of required modules and their clinical performance-based test and KOOS scores. Also, for the KOA group, Spearman’s rank correlation was used to determine whether the number of required modules and KL grade were correlated. A significance level of α<0.05 was set *a priori* for all statistical tests. All statistics analyses were performed in MATLAB 2016a (MathWorks, Natick, MA).

## RESULTS

### Population Average Demographics and Activation Patterns

Compared to the younger and older adults, the individuals with KOA had significantly greater mass (all *p* ≤ 0.002) and BMI (all *p* < 0.001) (Table 1). On average, the KOA group also walked significantly slower (*p* = 0.005) and with a shorter stride length (*p* = 0.010) than the younger adults. There was no statistically significant difference in age between the older adult and KOA groups (*p* = 0.946). As expected, the older adults and individuals with KOA were significantly older than the younger adults (all *p* < 0.001).

**Table 1:** Population Demographics

|  | Younger<br>(n = 10) | Older<br>(n = 10) | KOA<br>(n = 10) | p-Value |
| --- | --- | --- | --- | --- |
| Sex | 5M, 5F | 5M, 5F | 3M, 7F |  |
| Age (years) | 23.5 ± 3.1 | 63.5 ± 3.4 | 64.0 ± 4.0 | < 0.001<sup>a,b</sup> |
| Height (m) | 1.75 ± 0.07 | 1.68 ± 0.08 | 1.70 ± 0.10 | 0.209 |
| Mass (kg) | 71.2 ± 10.0 | 71.5 ± 13.5 | 89.7 ± 7.5 | < 0.001<sup>b,c</sup> |
| BMI (kg/m<sup>2</sup>) | 23.1 ± 2.8 | 24.8 ± 2.6 | 32.3 ± 4.1.9 | < 0.001<sup>b,c</sup> |
| Speed (m/s) | 1.24 ± 0.15 | 1.13 ± 0.20 | 0.95 ± 0.21 | 0.007<sup>b</sup> |
| Stride Length (Normalized by Leg Length) | 1.68 ± 0.17 | 1.51 ± 0.17 | 1.37 ± 0.28 | 0.013<sup>b</sup> |
| Number of Modules | 3.2 ± 0.4 | 2.7 ± 0.5 | 2.3 ± 0.7 | 0.004<sup>b</sup> |

Symbols indicate statistically significant pairwise differences between:
<sup>a</sup> Younger and Older
<sup>b</sup> Younger and KOA
<sup>c</sup> Older and KOA

The normalized EMG of the eight muscles exhibited similar activation patterns across populations (Figure 1). However, the normalized activation magnitude of the KOA group tended to be higher during stance compared to that of the older and younger adults across all muscles. Swing phase activation magnitudes were similar across all three groups.

**Figure 1:**
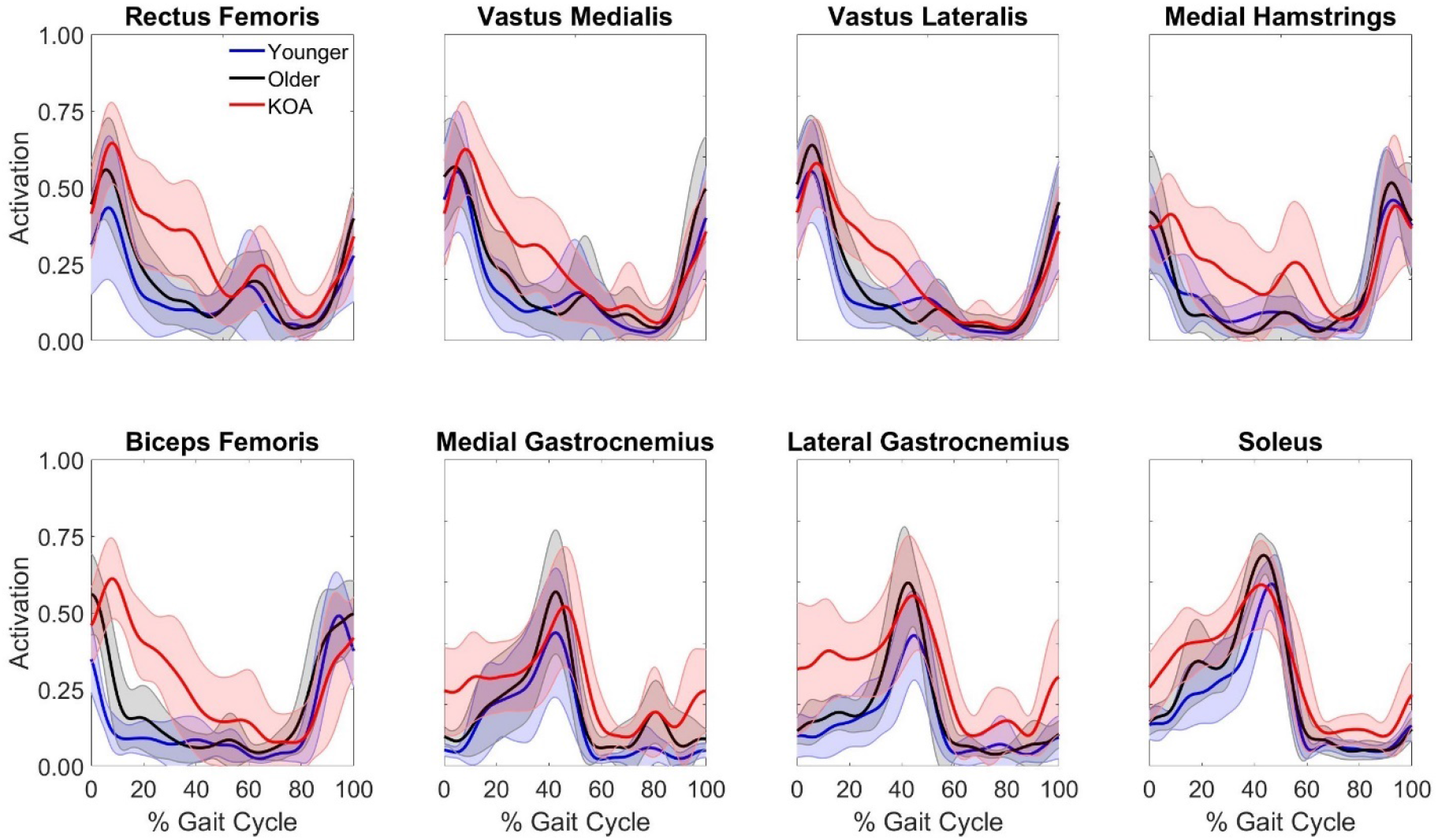
Average normalized EMG of young adults, older adults, and KOA patients. Solid lines represent population average. Shaded areas represent ± one standard deviation.

### Modular Control Complexity

Fewer modules were required to reconstruct the EMG of the KOA group (2.3 ± 0.7 modules) than the younger adult group (3.2 ± 0.4 modules) (*p* = 0.002; Table 1). The number of modules required for the older adults (2.7 ± 0.5 modules) did not significantly differ from the other two groups (*p* ≥ 0.113). Of the 10 younger adults, eight participants required three modules and two required four modules. Of the 10 older adults, three participants required two modules and seven required three modules. Of the 10 individuals with KOA, one participant required one module, five required two modules, and four required three modules. There were significant differences in minimum VAF between groups when one through seven modules were extracted by NMF (Figure 2; all *p* ≤ 0.005). The KOA group had significantly greater VAF than the younger and older adults when one module (both *p* < 0.001) and two modules (both *p* ≤ 0.014) were extracted. The KOA group had significantly greater VAF than the younger adults when three (*p* = 0.001), four (*p* = 0.001), five (*p* < 0.001), six (*p* = 0.002), and seven (*p* = 0.005) modules were extracted. The older adults had significantly greater VAF than the younger adults when four (*p* = 0.029), five (*p* = 0.007), and six (*p* = 0.025) modules were extracted.

**Figure 2:**
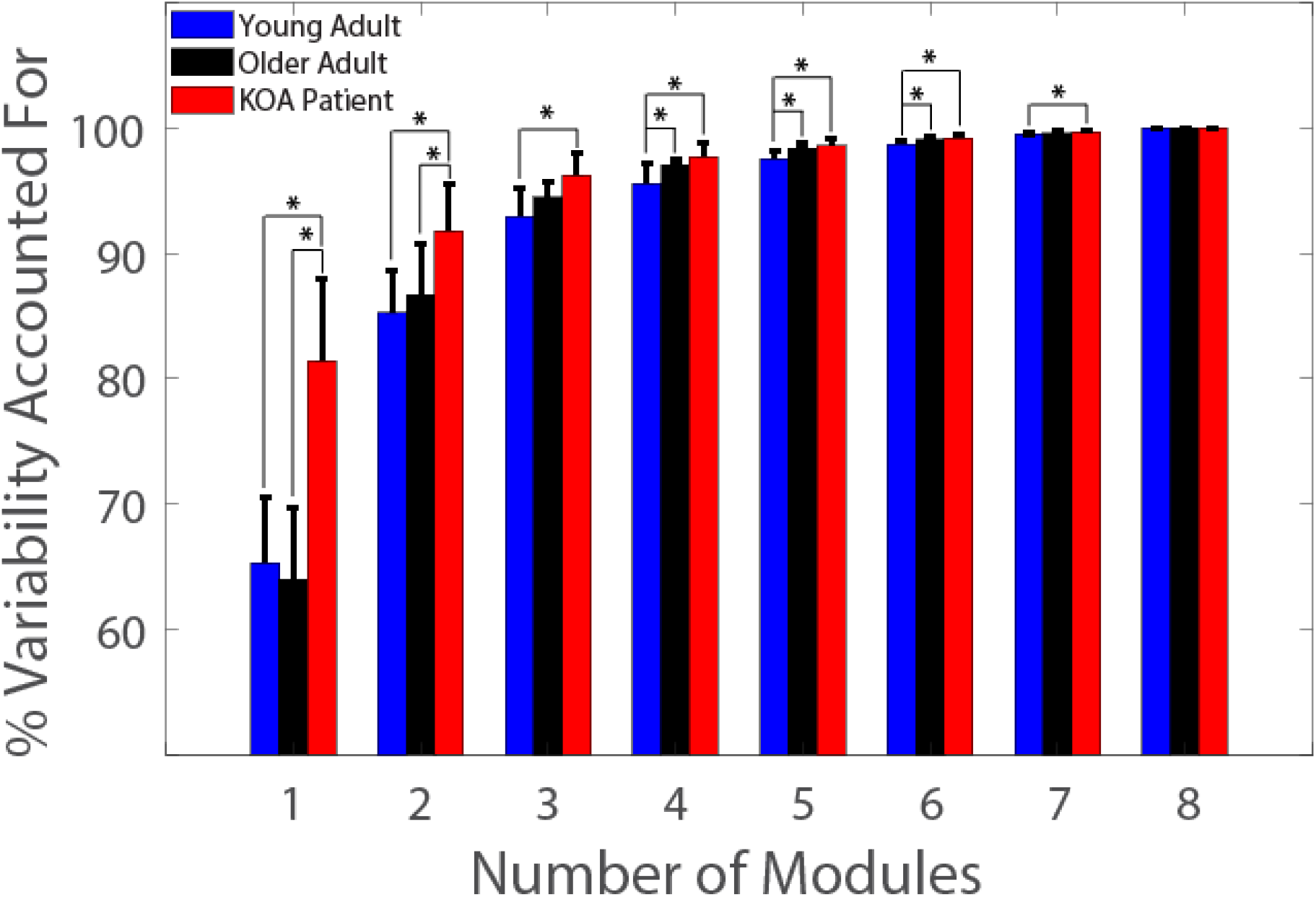
Total variability accounted for by *n* modules. * indicate significant pairwise differences between populations.

### Module Composition and Activation Timing

Although the total number of modules differed between subjects across the populations, four common modules were identified between at least two participants within two or more populations (Table 2; Figures 3-5): 1) the QUAD module characterized by high muscle weightings for the rectus femoris, vastus medialis, and vastus lateralis, 2) the HAMS module characterized by high muscle weightings for the medial hamstrings and biceps femoris, 3) the PF module characterized by high muscle weightings for the medial and lateral gastrocnemius and soleus, and 4) the QH module characterized by high muscle weightings for the rectus femoris, vastus medialis, vastus lateralis, medial hamstrings, and biceps femoris. There were strong correlations in the muscle weightings between all three populations for the QUAD (*r* ≥ 0.931), HAMS (*r* ≥ 0.895), and PF (*r* ≥ 0.996) modules (Table 2). The QH module was only observed in older adults (n = 3) and KOA participants (n=4) and there was a strong correlation in the QH muscle weightings between these populations (*r* = 0.990).

**Figure 3:**
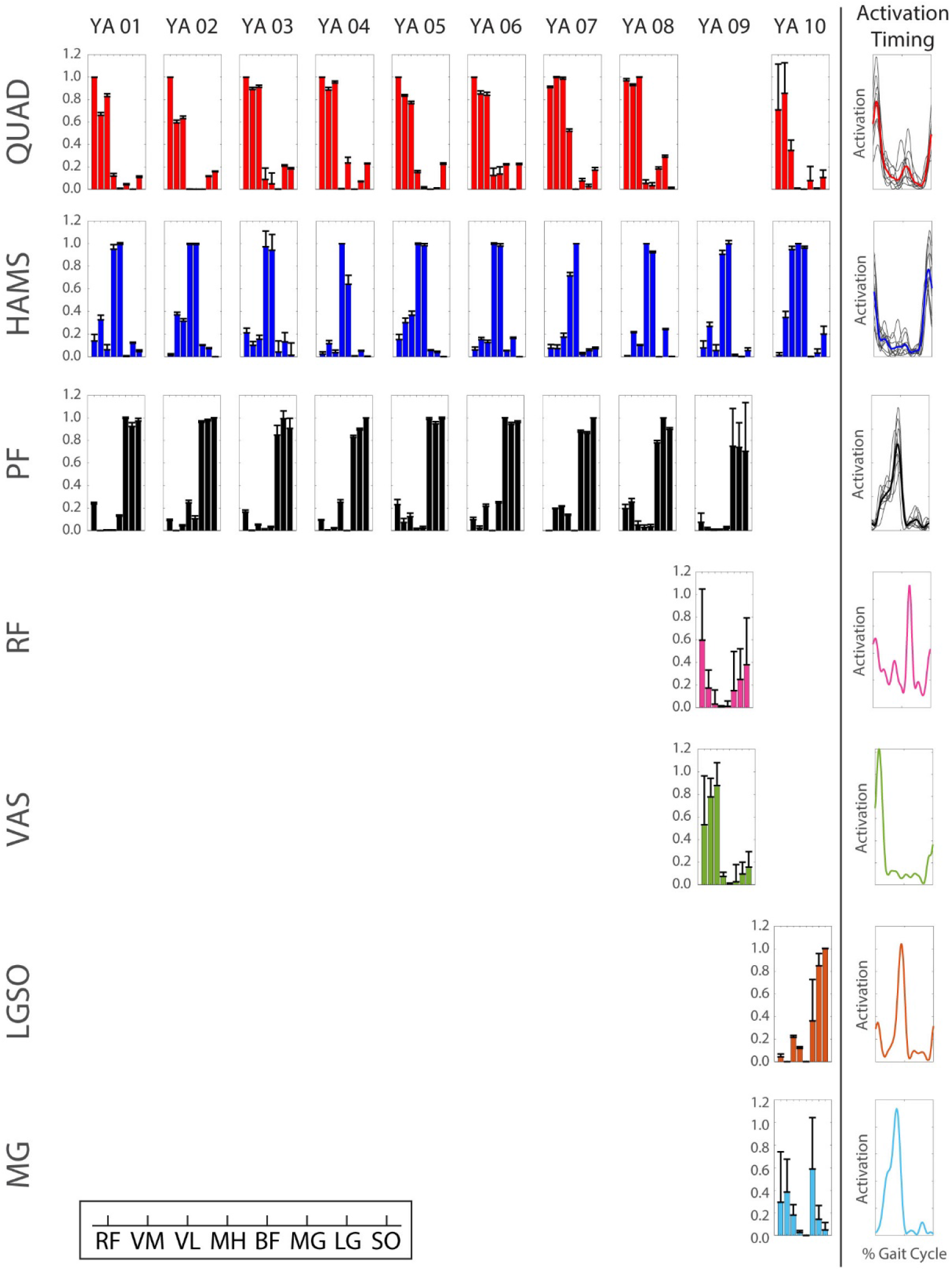
Young adult (YA) modular control. Bar heights represent the average bootstrapped muscle weights for the respective subject for the respective module. Error bars represent standard deviation of the bootstrapped weights. Bold, colored curves represent average activation timing profile across all subjects with the respective module. Gray curves represent individual subject activation timing profiles. Muscle abbreviations: rectus femoris (RF), vastus medialis (VM), vastus lateralis (VL), medial hamstrings (MH), biceps femoris (BF), medial gastrocnemius (MG), lateral gastrocnemius (LG), soleus (SO).

**Figure 4:**
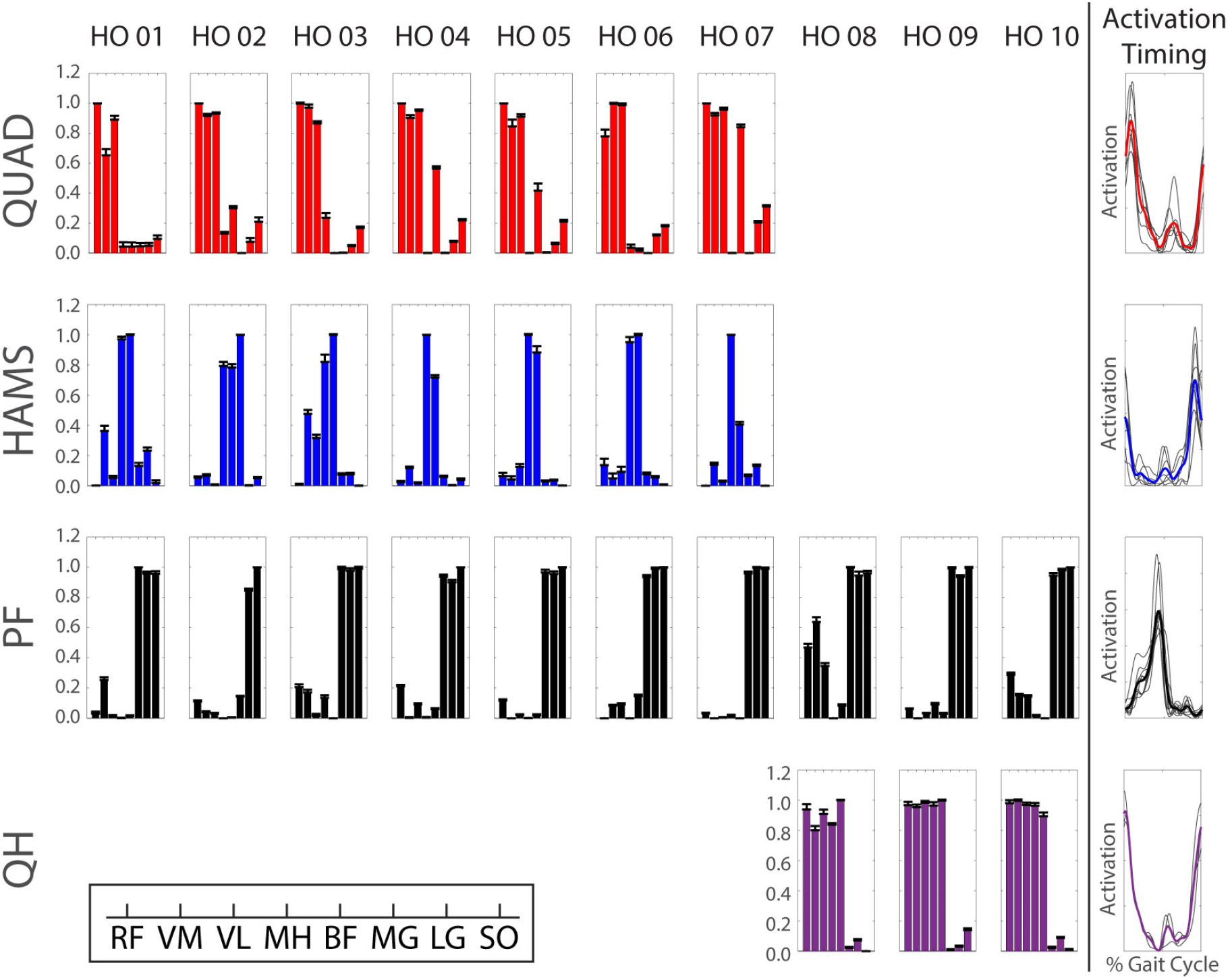
Healthy older (HO) adult modular control. Bar heights represent the average bootstrapped muscle weights for the respective subject for the respective module. Error bars represent standard deviation of the bootstrapped weights. Bold, colored curves represent average activation timing profile across all subjects with the respective module. Gray curves represent individual subject activation timing profiles. Muscle abbreviations: rectus femoris (RF), vastus medialis (VM), vastus lateralis (VL), medial hamstrings (MH), biceps femoris (BF), medial gastrocnemius (MG), lateral gastrocnemius (LG), soleus (SO).

**Figure 5:**
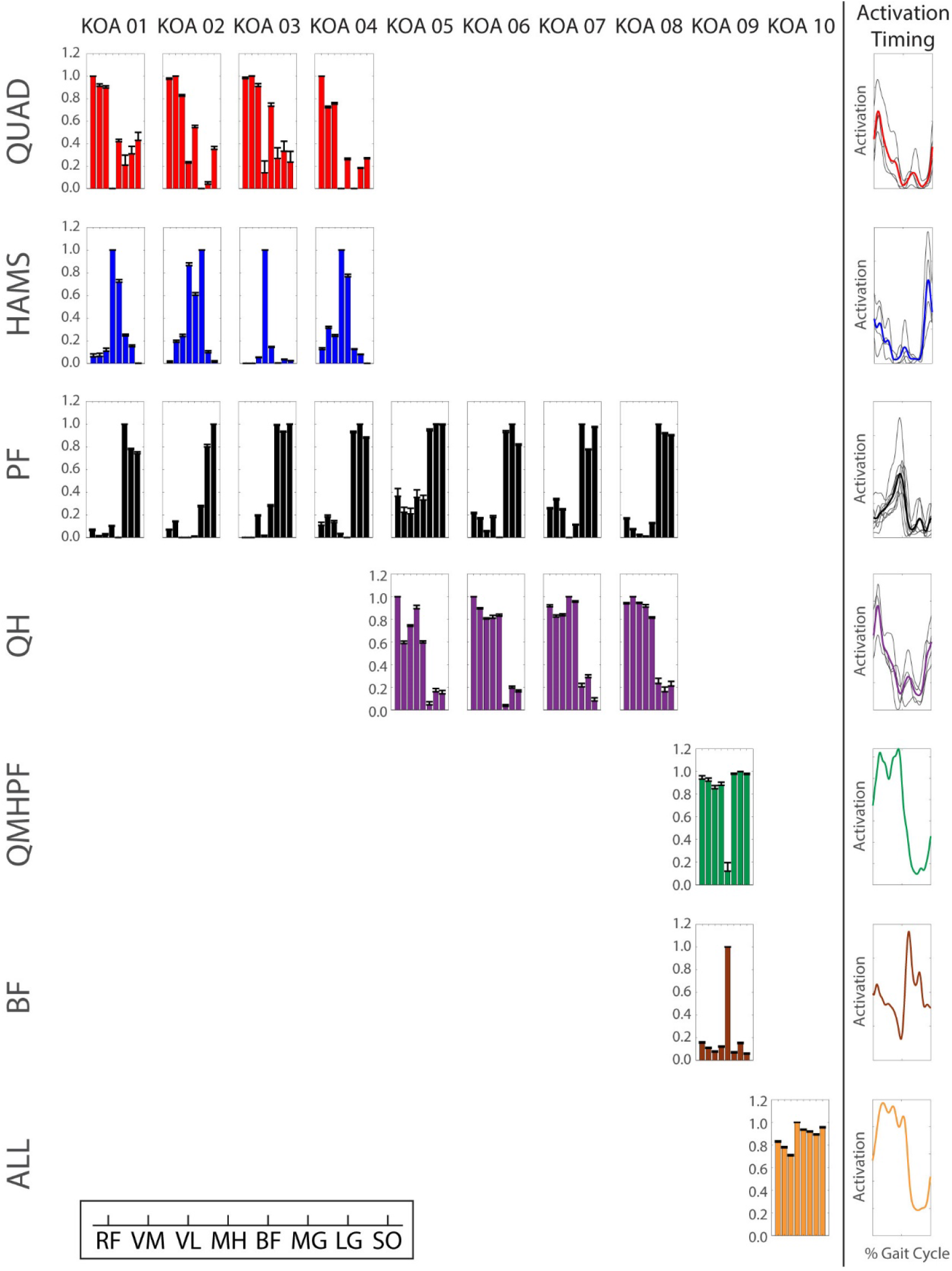
KOA patient modular control. Bar heights represent the average bootstrapped muscle weights for the respective subject for the respective module. Error bars represent standard deviation of the bootstrapped weights. Bold, colored curves represent average activation timing profile across all subjects with the respective module. Gray curves represent individual subject activation timing profiles. Muscle abbreviations: rectus femoris (RF), vastus medialis (VM), vastus lateralis (VL), medial hamstrings (MH), biceps femoris (BF), medial gastrocnemius (MG), lateral gastrocnemius (LG), soleus (SO).

**Table 2:**
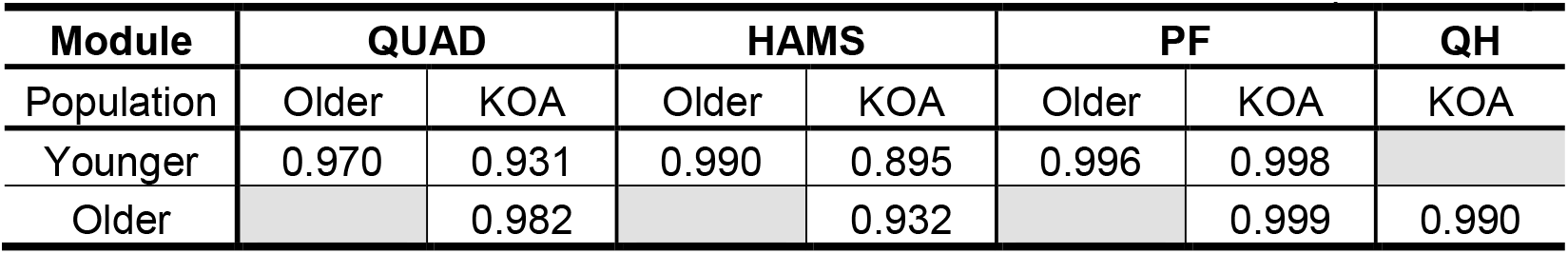
Common Module Pearson Correlation Coefficients Between Populations

| Module | QUAD |  | HAMS |  | PF |  | QH |
| --- | --- | --- | --- | --- | --- | --- | --- |
| Population | Older | KOA | Older | KOA | Older | KOA | KOA |
| Younger | 0.970 | 0.931 | 0.990 | 0.895 | 0.996 | 0.998 |  |
| Older |  | 0.982 |  | 0.932 |  | 0.999 | 0.990 |

Within the group, common modules were generally correlated across all participants who were classified as having the module (Table 3). In the young adults, the common modules were correlated between all participants with that common module. In the older adults, the QUAD modules of all seven individuals with a QUAD module were correlated and the QH modules of all three individuals with a QH module were correlated. However, the HAMS and PF modules of one older adult (healthy older (HO) adult 02) were not correlated with those of the other six and nine older adults who had a HAMS and PF module, respectively. In addition, the HAMS module of one individual with KOA (KOA 02) did not correlate with the HAMS module of the other three individuals with KOA who had a HAMS module. Otherwise, the common modules of the individuals with KOA were correlated between all individuals with that common module.

**Table 3:** Number of Participants with Correlated Module within Population*

| Population | # of Modules** | QUAD Module | HAMS Module | PF Module | QH Module |
| --- | --- | --- | --- | --- | --- |
| Younger | 3 (n=8) | 8/8 | 8/8 | 8/8 | 0/0 |
|  | 4 (n=2) | 1/1 | 2/2 | 1/1 | 0/0 |
| Older | 2 (n=3) | 0/0 | 0/0 | 3/3 | 3/3 |
|  | 3 (n=7) | 7/7 | 6/7 | 6/7 | 0/0 |
| KOA | 1 (n=1) | 0/0 | 0/0 | 0/0 | 0/0 |
|  | 2 (n=5) | 0/0 | 0/0 | 4/4 | 4/4 |
|  | 3 (n=4) | 4/4 | 3/4 | 4/4 | 0/0 |

\* Number of participants within a population with correlated modules / total number of participants within the population classified by K-means clustering to have that module
\*\* n = number of participants within the population that have X number of modules

There were no differences in W_musc_ between groups for any of the four common modules (Table 4; all *p* ≥ 0.485). However, the W_sum_ of the QUAD module was greater in the individual KOA than in the younger adults when all participants with the QUAD module were considered, regardless of modular control complexity (*p* = 0.025). When only individuals with 3 modules and a QUAD module were included in the analysis (excludes one younger adult with 4 modules), the individuals KOA still had a greater W_sum_ than the younger adults (*p* = 0.040).

**Table 4:** W_musc_ and W_sum_ for Common Modules

| Module | Population | $W_{\text{musc}}$ | $W_{\text{sum}}$ |
| --- | --- | --- | --- |
| **QUAD** | Younger (n=9) | $7.7 \pm 0.5$ | $3.1 \pm 0.5$ |
| | Older (n=7) | $7.9 \pm 0.4$ | $3.5 \pm 0.5$ |
| | KOA (n=4) | $7.5 \pm 0.6$ | $4.0 \pm 0.6^*$ |
| **HAMS** | Younger (n=10) | $7.9 \pm 0.3$ | $2.6 \pm 0.4$ |
| | Older (n=6) | $7.8 \pm 0.4$ | $2.3 \pm 0.4$ |
| | KOA (n=3) | $7.7 \pm 0.6$ | $2.1 \pm 0.7$ |
| **PF** | Younger (n=9) | $7.7 \pm 0.5$ | $3.2 \pm 0.4$ |
| | Older (n=9) | $7.6 \pm 0.7$ | $3.4 \pm 0.4$ |
| | KOA (n=8) | $7.5 \pm 0.5$ | $3.3 \pm 0.6$ |
| **QH** | Older (n=3) | $8.0 \pm 0.0$ | $4.8 \pm 0.2$ |
| | KOA (n=4) | $8.0 \pm 0.0$ | $4.8 \pm 0.5$ |

\* significantly different from Younger

Module activation timing profiles differed between at least two groups for each of the four common modules (Figure 6). The older adults had greater AUC in the QUAD module during late swing than the KOA group (*p* = 0.018). The older adults had greater AUC in the HAMS module during early swing than the younger adults (*p* = 0.043). In the PF module, the older adults had a greater late midstance AUC (*p* = 0.006), but a smaller late swing AUC than the KOA group (*p* = 0.001). The younger adults also had a smaller late swing AUC than the participants KOA (*p* < 0.001). In the QH module the older adults had a greater weight acceptance AUC (*p* = 0.043), but smaller early (*p* = 0.030) and late (*p* = 0.035) midstance AUCs than the individuals with KOA. There were no significant differences in AUC for all other gait phases in the common modules (*p* ≥ 0.050).

**Figure 6:**
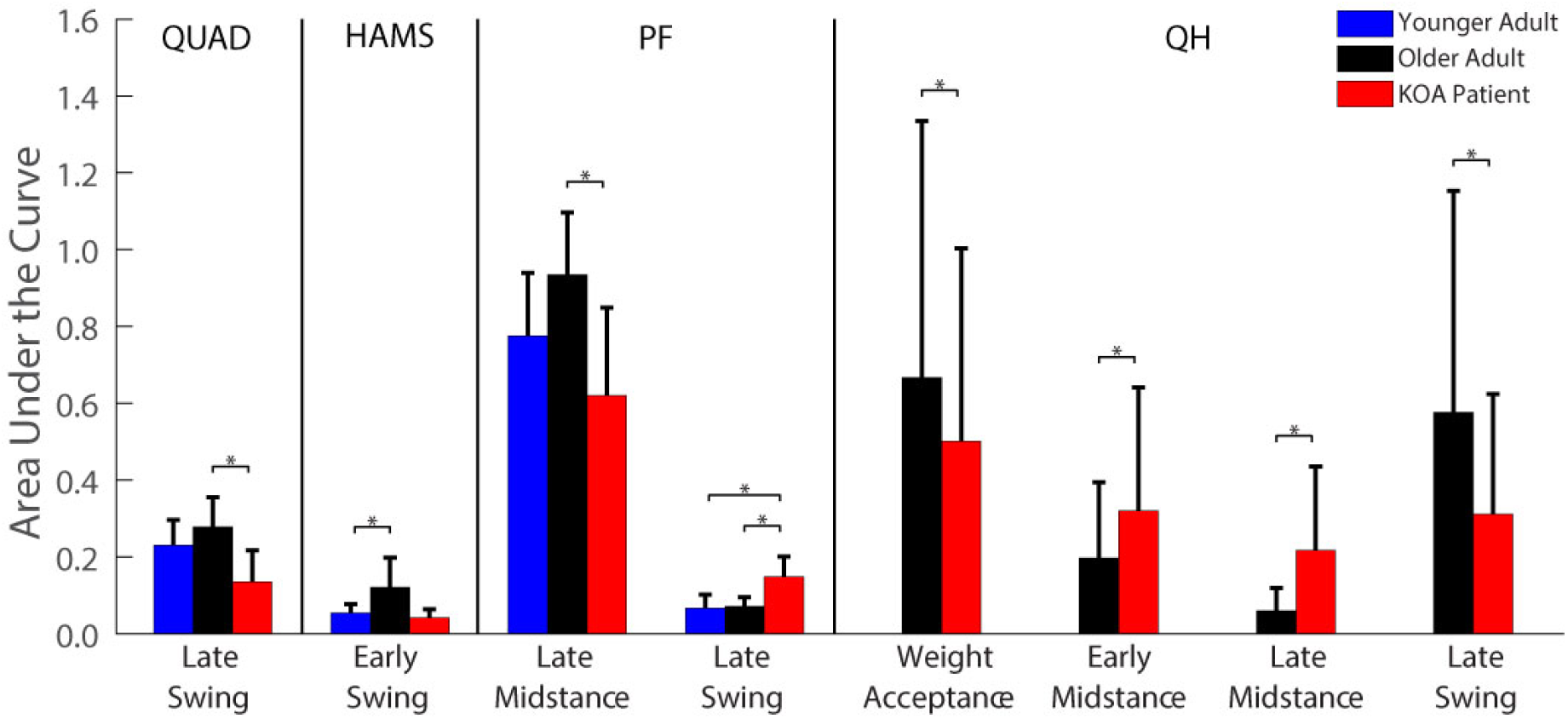
Area under the curve for common module activation timing profiles with significant differences between populations.

### Joint Kinematics and Kinetics

There were several differences in peak joint kinematics and kinetics between populations (Table 5; Figure 7). Compared to the older adult and KOA groups, the younger adults had smaller peak hip flexion angles in early stance (H1: *p* ≤ 0.001) and swing (H3: *p* ≤ 0.018), greater hip extension (H2: *p* ≤ 0.006), less knee flexion in early stance (K1: *p* ≤ 0.015), greater knee extension (K2: *p* ≤ 0.001), and less dorsiflexion during swing (A4: *p* ≤ 0.005). The older adults had a greater peak knee flexion angle during late stance than the younger adults and individuals with KOA (K3: *p* ≤ 0.027). The KOA group had a smaller peak hip extension angle than the older adults (H2: *p* = 0.014) and a greater peak late stance dorsiflexion angle (A2: *p* = 0.007) and a smaller peak plantarflexion angle (A3: *p* = 0.014) than the younger adults. With respect to joint kinetics, the younger adults had a greater hip flexion moment than the older adults and KOA participants (HM2: *p* ≤ 0.018). The younger adults also had a greater midstance peak knee flexion moment (KM3: *p* = 0.001) than the individuals with KOA. Compared to the younger adults, the older adults had a smaller peak weight acceptance knee flexion moment (KM1: *p* = 0.018) and a greater early stance peak knee extension moment (KM2: *p* = 0.036). There were no pairwise differences between populations for all other peak joint angles and moments (*p* ≥ 0.060).

**Table 5:** Peak Joint Angles and Moments

| Metric | Joint | Peak | Younger<br>(n = 10) | Older<br>(n = 10) | KOA<br>(n = 10) | p-Value |
| --- | --- | --- | --- | --- | --- | --- |
| Angle (°) | Hip | H1 | 24.7 ± 4.4 | 39.2 ± 5.7 | 39.5 ± 7.0 | <0.001<sup>a,b</sup> |
|  |  | H2 | -11.5 ± 4.8 | -1.3 ± 4.1 | 7.7 ± 9.6 | <0.001<sup>a,b,c</sup> |
|  |  | H3 | 31.4 ± 6.0 | 43.2 ± 4.5 | 40.2 ± 8.9 | 0.002<sup>a,b</sup> |
|  | Knee | K1 | 9.6 ± 8.3 | 24.5 ± 7.2 | 24.4 ± 15.4 | 0.006<sup>a,b</sup> |
|  |  | K2 | -1.8 ± 5.5 | 11.2 ± 3.8 | 14.5 ± 8.0 | <0.001<sup>a,b</sup> |
|  |  | K3 | 62.8 ± 3.8 | 74.7 ± 3.6 | 57.7 ± 15.8 | 0.002<sup>a,c</sup> |
|  | Ankle | A1 | -9.4 ± 3.8 | -5.4 ± 3.8 | -6.8 ± 4.6 | 0.105 |
|  |  | A2 | 4.6 ± 5.3 | 9.7 ± 3.8 | 12.4 ± 6.3 | 0.008<sup>b</sup> |
|  |  | A3 | -25.2 ± 9.3 | -17.7 ± 7.6 | -13.9 ± 7.8 | 0.017<sup>b</sup> |
|  |  | A4 | -2.0 ± 2.7 | 2.8 ± 3.5 | 4.5 ± 2.9 | <0.001<sup>a,b</sup> |
| Moment<br>(x10<sup>-2</sup> Nm/Nm*m/s<sup>-1</sup>) | Hip | HM1 | 3.7 ± 0.9 | 3.8 ± 0.9 | 3.8 ± 2.3 | 0.984 |
|  |  | HM2 | -4.2 ± 0.9 | -2.9 ± 0.7 | -2.9 ± 1.0 | 0.004<sup>a,b</sup> |
|  |  | HM3 | 1.4 ± 0.3 | 0.9 ± 0.5 | 1.4 ± 0.6 | 0.093 |
|  | Knee | KM1 | -1.7 ± 0.8 | -0.7 ± 0.7 | -1.4 ± 0.9 | 0.023<sup>a</sup> |
|  |  | KM2 | 1.1 ± 1.5 | 2.7 ± 1.0 | 2.3 ± 1.6 | 0.045<sup>a</sup> |
|  |  | KM3 | -1.6 ± 0.6 | -0.6 ± 0.5 | 0.3 ± 1.5 | 0.002<sup>b</sup> |
|  |  | KM4 | 1.1 ± 0.4 | 1.7 ± 0.5 | 1.4 ± 0.9 | 0.041 |
|  |  | KM5 | -1.1 ± 0.2 | -1.3 ± 0.4 | -1.0 ± 0.3 | 0.086 |
|  | Ankle | AM1 | -1.2 ± 0.3 | -1.0 ± 0.4 | -0.9 ± 0.4 | 0.357 |
|  |  | AM2 | 6.2 ± 1.0 | 6.6 ± 1.2 | 6.0 ± 2.2 | 0.620 |

Peak joint angles and moments reported in the table as the average ± standard deviation. Hip flexion, knee flexion, and ankle dorsiflexion *angles* are positive. Hip extension, knee extension, and ankle plantarflexion *moments* are positive and were normalized to body mass, height, and walking speed. Superscripted letters indicate pairwise differences between:
<sup>a</sup> Younger and Older
<sup>b</sup> Younger and KOA
<sup>c</sup> Older and KOA

**Figure 7:**
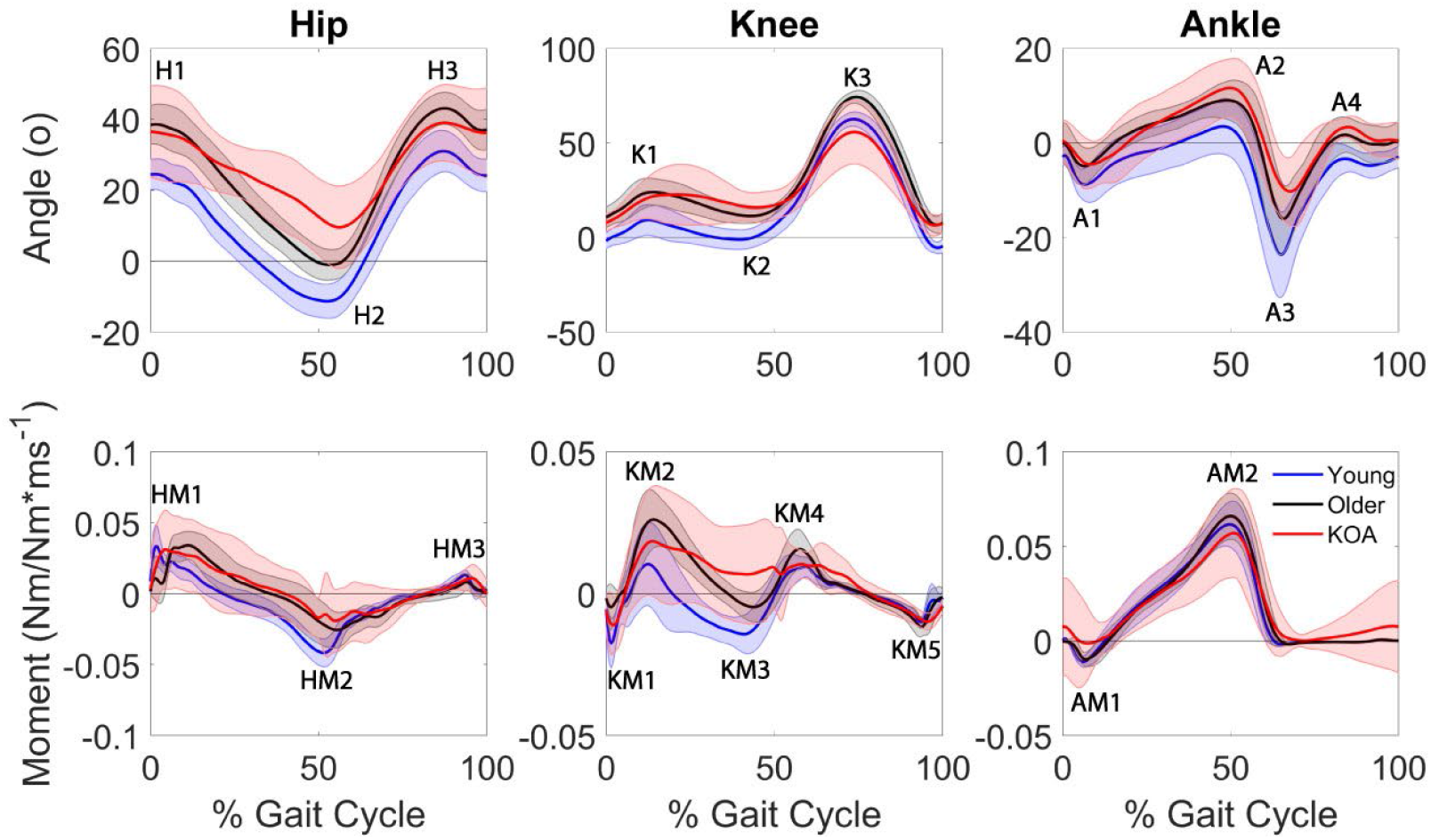
Population sagittal plane joint angles and joint moments (normalized to body weight and height). Solid lines represent population average and shaded error bars represent ± one standard deviation. Positive values represent hip flexion, knee flexion, and ankle dorsiflexion angles and hip extension, knee extension, and ankle plantarflexion moments.

The merging of the HAMS module with another module or additional muscles affected peak joint angles and moments (Table 6). Individuals demonstrating a HAMS module that was merged with other muscles had greater hip flexion during early stance (H1; *p* = 0.039) and swing phase (H3; *p* = 0.038) and greater late stance dorsiflexion (A2; *p* = 0.048) than individuals not having a merged HAMS module. A merged HAMS module also resulted in a smaller knee flexion moment during midstance (K3; *p* =0.030). A merged QUAD or PF module did not significantly alter peak joint angles or moments (*p* ≥ 0.057).

**Table 6:** Peak Joint Angles and Moments by Common Module

| Metric | Joint | Peak | QUAD Module |  | HAMS Module |  | PF Module |  |
| --- | --- | --- | --- | --- | --- | --- | --- | --- |
|  |  |  | Independent<br>(n=20) | Merged<br>(n=9) | Independent<br>(n=19) | Merged<br>(n=11) | Independent<br>(n=26) | Merged<br>(n=2) |
| Angle (°) | Hip | H1 | 33.1 ± 9.1 | 39.5 ± 5.3 | 31.9 ± 9.9 | 38.9 ± 5.0* | 34.1 ± 9.2 | 39.3 ± 8.5 |
|  |  | H2 | -3.4 ± 9.2 | 3.7 ± 10.6 | -4.4 ± 9.8 | 3.0 ± 9.7 | -2.6 ± 9.1 | 8.9 ± 16.5 |
|  |  | H3 | 37.2 ± 9.0 | 42.1 ± 4.2 | 35.9 ± 9.3 | 42.3 ± 3.9* | 37.8 ± 8.6 | 42.5 ± 5.8 |
|  | Knee | K1 | 21.4 ± 14.3 | 17.5 ± 6.5 | 20.6 ± 15.4 | 17.6 ± 5.9 | 20.8 ± 12.5 | 17.3 ± 2.6 |
|  |  | K2 | 6.8 ± 10.2 | 11.5 ± 5.9 | 6.2 ± 10.6 | 11.0 ± 5.5 | 8.2 ± 9.0 | 12.3 ± 4.4 |
|  |  | K3 | 66.0 ± 13.1 | 63.7 ± 9.2 | 65.3 ± 13.4 | 64.7 ± 8.8 | 65.6 ± 12.2 | 61.8 ± 11.3 |
|  | Ankle | A1 | -7.6 ± 4.2 | -6.1 ± 4.7 | -7.8 ± 4.3 | -6.2 ± 4.3 | -6.7 ± 3.7 | -7.5 ± 4.7 |
|  |  | A2 | 7.6 ± 6.4 | 12.2 ± 3.9 | 7.2 ± 6.3 | 11.7 ± 4.3* | 8.8 ± 6.3 | 10.2 ± 5.5 |
|  |  | A3 | -20.9 ± 9.2 | -15.7 ± 8.9 | -19.8 ± 9.6 | -17.5 ± 8.9 | -19.0 ± 8.9 | -13.8 ± 11.1 |
|  |  | A4 | 1.2 ± 4.5 | 3.5 ± 2.8 | 0.9 ± 4.4 | 3.2 ± 3.0 | 2.0 ± 4.0 | 2.0 ± 3.2 |
| Moment<br>(x10<sup>-2</sup> Nm/Nm*m/s<sup>-1</sup>) | Hip | HM1 | 3.7 ± 1.5 | 3.9 ± 1.6 | 3.8 ± 1.5 | 3.7 ± 1.5 | 3.7 ± 1.3 | 4.2 ± 3.0 |
|  |  | HM2 | -3.3 ± 1.1 | -3.2 ± 0.8 | -3.4 ± 1.3 | -3.3 ± 0.7 | -3.3 ± 1.1 | -4.2 ± 0.4 |
|  |  | HM3 | 1.2 ± 0.6 | 1.3 ± 0.5 | 1.1 ± 0.5 | 1.4 ± 0.6 | 1.2 ± 0.6 | 1.4 ± 0.5 |
|  | Knee | KM1 | -1.3 ± 0.9 | -1.3 ± 0.9 | -1.3 ± 0.9 | -1.3 ± 0.9 | -1.2 ± 0.8 | -1.6 ± 1.7 |
|  |  | KM2 | 2.2 ± 1.4 | 1.9 ± 1.5 | 2.0 ± 1.6 | 2.1 ± 1.4 | 2.1 ± 1.4 | 2.4 ± 1.7 |
|  |  | KM3 | -0.9 ± 1.0 | 0.0 ± 1.5 | -1.0 ± 1.0 | 0.0 ± 1.3* | -0.7 ± 1.2 | 0.5 ± 0.7 |
|  |  | KM4 | 1.5 ± 0.7 | 1.6 ± 0.7 | 1.5 ± 0.7 | 1.5 ± 0.7 | 1.5 ± 0.6 | 2.0 ± 0.9 |
|  |  | KM5 | -1.2 ± 0.4 | -1.1 ± 0.2 | -1.1 ± 0.4 | -1.2 ± 0.2 | -1.1 ± 0.3 | -1.3 ± 0.2 |
|  | Ankle | AM1 | -1.1 ± 0.4 | -0.9 ± 0.3 | -1.1 ± 0.4 | -0.9 ± 0.3 | -1.0 ± 0.4 | -0.9 ± 0.3 |
|  |  | AM2 | 6.1 ± 1.6 | 6.5 ± 1.4 | 6.2 ± 1.7 | 6.4 ± 1.3 | 6.2 ± 1.5 | 7.4 ± 1.8 |

Peak joint angles and moments reported as the average ± standard deviation. Hip flexion, knee flexion, and ankle dorsiflexion *angles* are positive. Hip extension, knee extension, and ankle plantarflexion *moments* are positive and were normalized to body mass, height, and walking speed. \* indicates statistical differences due to the absence of a common module.

### Associations between Module Number and Clinical and Performance Metrics

Self-selected walking speed was positively associated with the number of required modules when all participants were grouped together such that individuals who walked faster required a greater number of modules (*τ*_*b*_ = 0.350, *p* = 0.021; Table 7). However, within a population, only the KOA group had a significant positive correlation between the number of required modules and self-selected walking speed (*τ*_*b*_ = 0.581, *p* = 0.045). There were no signficant associations between the number of required modules and self-selected walking speed for the younger or older adults (*p* ≥ 0.649).

**Table 7:** Self-Selected Walking Speed* (m/s) By Population and Number of Modules

| Population | # of Modules |  |  |  | By Population |  | All Participants |  |
| --- | --- | --- | --- | --- | --- | --- | --- | --- |
| | 1 | 2 | 3 | 4 | $\tau_b$ | $p$-value | $\tau_b$ | $p$-value |
| YA | | | 1.23 ± 0.14 | 1.30 ± 0.28 | 0.113 | 0.793 | | |
| OA | | 1.18 ± 0.16 | 1.11 ± 0.22 | | -0.163 | 0.649 | 0.350 | 0.021<sup>a</sup> |
| KOA | 0.56 ± 0.00 | 0.93 ± 0.17 | 1.08 ± 0.11 | | 0.581 | 0.045<sup>a</sup> | | |

\* Speed reported as average ± standard deviation. <sup>a</sup> significant correlation.

There was a significant association between the number of modules required by the KOA group and the ADL subscale of the KOOS (*τ*_*b*_ = 0.644, *p* = 0.021) with higher (better) scores associated with a greater number of modules (Table 8). There were no significant correlations between the number of required modules and other KOOS subscales (all *p* ≥ 0.055), KL grade (*p* = 0.316), or the performance-based clinical assessments (all *p* ≥ 0.133).

**Table 8:** Knee Osteoarthritis Clinical Assessments by Module Group

| # of Modules | KL Grade | Performance-Based Outcomes |  |  | KOOS Scores |  |  |  |
| --- | --- | --- | --- | --- | --- | --- | --- | --- |
|  |  | 6MW (m) | SCT (s) | TUG (s) | Pain | Symptoms | ADL* | QOL |
| One Modules (n=1) | 4.0 ± 0.0 | 180.4 ± 0.0 | 39.9 ± 0.0 | 15.4 ± 0.0 | 39.0 ± 0.0 | 50.0 ± 0.0 | 40.0 ± 0.0 | 25.0 ± 0.0 |
| Two Modules (n=5) | 3.0 ± 0.0 | 379.4 ± 84.8 | 33.7 ± 17.4 | 10.9 ± 2.9 | 42.0 ± 12.1 | 44.4 ± 17.8 | 53.2 ± 11.0 | 22.8 ± 18.1 |
| Three Modules (n=4) | 3.8 ± 0.5 | 434.0 ± 80.5 | 18.3 ± 14.7 | 9.4 ± 0.4 | 74.3 ± 21.3 | 67.8 ± 20.7 | 75.5 ± 18.3 | 47.0 ± 28.5 |

\* Significant positive association with number of modules

## DISCUSSION

This pilot study aimed to determine whether age and KOA alter the modular control of walking. The experimental activation patterns of all participants were able to be reduced to a modular organization. Thus, the findings of this study contribute to the evidence for a low-dimensional organization of the neural control of muscles during gait across age groups and in the presence of pathology. Individuals with KOA required fewer modules to adequately reconstruct their experimental activation patterns than younger adults and had a greater VAF than both younger and older adults when just one and two modules were extracted and a greater VAF than the younger adults when 3-7 modules were extracted. Although the older adults did have marginally (on average, 0.4-1.4%) greater VAF than the younger adults when 4-6 modules were extracted, which indicates a slightly decrease in variability in the muscle activation patterns due to age, the number of modules required by the older adults did not significantly differ from the other two populations. These findings suggest aging alone may not significantly reduce modular control complexity. However, the combined effects of knee osteoarthritis and advanced age may together reduce modular control complexity.

The number of required modules in each population in this study (younger: 3-4, older: 2-3, KOA: 1-3) was generally consistent with previous analyses of modular control during walking in healthy younger and older adults and patient populations (e.g., Allen et al. 2019; Allen and Franz 2018; Clark et al. 2010b, Ardestani et al. 2017). The selection of muscles included in a module analysis influences the resulting modules (51). Thus, with the muscle set collected in this study comprised of three major muscle groups (the quadriceps, hamstrings, and plantarflexors), it is reasonable that three modules are sufficient for independent control of these muscles. Indeed, 19 of the 30 total participants required three modules. Only two younger adults required more than three modules, which may provide greater flexibility in an individual’s control strategy. For example, independent control of the biarticular rectus femoris and uniarticular vasti (Figure 3: young adult (YA) 09) would promote greater independence of the hip flexor and knee extensor actions. Most participants with two modules exhibited merged quadriceps and hamstrings activation, with the exception of one KOA participant who exhibited merged activation of all muscles except for the biceps femoris (Figure 5: KOA 09). One KOA patient only required one module to adequately reconstruct the activation patterns of all eight muscles (Figure 5: KOA 10). This individual exhibited a relatively constant activation of all eight EMG muscles throughout the stance phase (Figure 8), which is indicative of a very simplistic “all or nothing” control strategy that activates all or none of the muscles at a given time.

**Figure 8:**
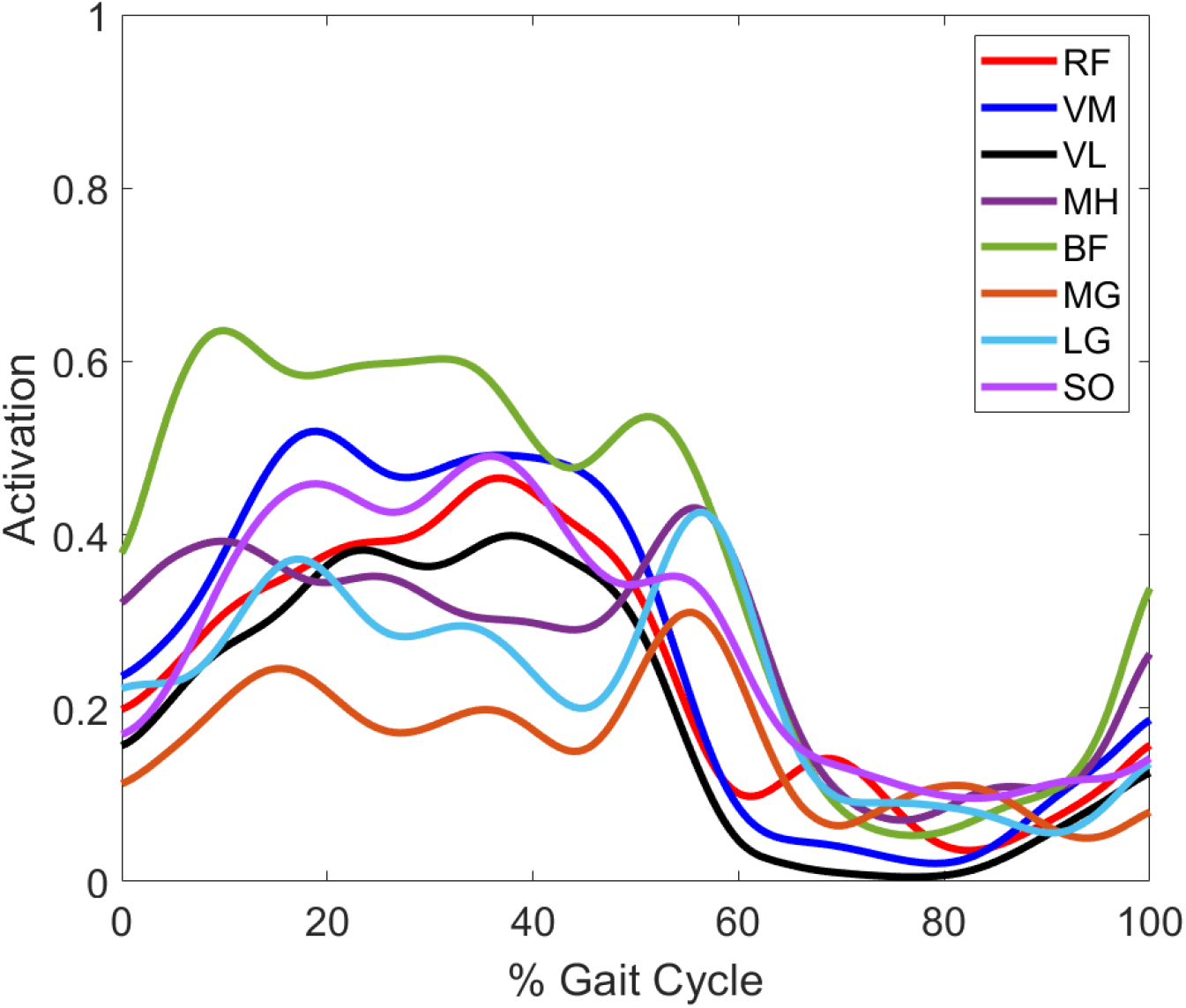
Average muscle EMG for the KOA participant with one module (KOA 10). Muscle abbreviations: rectus femoris (RF), vastus medialis (VM), vastus lateralis (VL), medial hamstrings (MH), biceps femoris (BF), medial gastrocnemius (MG), lateral gastrocnemius (LG), soleus (SO).

The three common modules identified in the younger adults (i.e., QUAD, HAMS, and PF) were also observed in a majority of the older adults. For those younger and older adults with the common modules, the module compositions were very strongly correlated between age groups (*r* ≥ 0.970). The older adults’ HAMS module did exhibit greater area under the curve during early swing than that of the young adults, which may have contributed to the older adults’ increased knee flexion during swing due to earlier hamstrings activation in the older adults than in the younger adults (or individuals with KOA). However, W_sum_ did not differ between the healthy younger and older adults for any of the common modules, which indicates that, when those independent modules are present, muscle co-activity does not increase significantly. Together, these findings suggest the nervous system may be capable of preserving modular control with unimpaired aging.

It is important to note that there was no statistically significant difference in walking speed between the younger and older adults in this study, which suggests the older adults in this study had a relatively high level of function. Although consistent modular organization has been observed across walking speeds in healthy younger and older adults (22,23), the number of required modules was significantly associated with self-selected walking speed when all participants in this study were considered (Table 7). In addition, previous studies have reported an association between slower walking speeds and altered modular organization in patient populations (23,49). Therefore, future studies should investigate modular control in older adults with slower self-selected walking speeds to clarify the relationship between age-related changes in motor control and functional performance.

The KOA group exhibited altered module composition and timing compared to the unimpaired younger and older adults, which suggests these differences in modular control may be disease-related. Only four of the 10 participants with KOA had independent QUAD and HAMS modules (compared to seven of the 10 older adults). The other six participants with KOA exhibited merging of the quadriceps muscles with the hamstrings and/or plantarflexors. Those individuals with KOA with an independent QUAD module had a greater W_sum_ than the younger adults, which suggests that even when the QUAD module is independent in the KOA group (i.e., not merged with the HAMS module or other muscles), there is still an increase in co-activity of other muscles with the quadriceps muscles. Compared to the three older adults with a QH module, the four KOA participants with a QH module had greater module activation during early and late midstance, the phases of the gait cycle when the affected leg is primarily responsible for supporting the body. This is in contrast to the smaller QH module activation observed in the KOA group during late swing and weight acceptance when they would have been fully or partially supported by the contralateral limb. The increased QH activation during midstance observed in the KOA group but not the unimpaired older adults may be a disease-related control strategy to stabilize the knee during single-leg support. Thus, although the QH module composition was very strongly correlated between the older adults and KOA group (*r* = 0.990), the module activation pattern was significantly influenced by the presence of KOA.

Notably, the number of significantly active muscles per module (W_musc_) did not differ between populations for any of the common modules. The average W_musc_ was greater than or equal to 7.5 for all populations and all common modules, indicating that the weight of most or all muscles was significantly different from 0 and, therefore, statistically associated with the module. However, visual inspection of the muscle weights for the common modules for each population in Figures 3-5 shows that the contributions outside of the primary muscles associated with the module were generally minimal. A clear exception is the QUAD module of the KOA group, for which multiple muscles in addition to the quadriceps had large weights, which is consistent with their larger W_sum_.

The absence of the HAMS module resulted in greater peak hip flexion and ankle dorsiflexion angles and a smaller peak midstance knee flexion moment. The KOA groups’ prolonged knee extension moment during midstance compared to the flexion moment observed in the younger adults is likely related to the high prevalence of merging of the control of the hamstring muscles with the quadriceps and plantarflexors in the individuals with KOA. The increased midstance activation of the QH module (when the HAMS module is merged with the QUAD module), along with the larger W_sum_ when the QUAD module was present in the participants with KOA, likely contribute to the KOA groups’ midstance knee extension moment. Thus, merging of typically independent modules can alter joint moment production ability, likely due to the inappropriately timed activation of antagonist muscles.

The altered neuromuscular control strategy identified in the KOA group in this study suggests that the reduced modular control complexity observed in individuals who had undergone a TKA (35) may have been present prior to surgery. Thus, replacement of the joint structure alone may not be sufficient to improve locomotor performance in individuals with end-stage KOA. However, additional research is required to determine if pre-operative modular control is associated with post-operative modular control. Retraining the neuromuscular control strategy may be required to achieve improvements in walking speed and coordination in individuals with KOA or following a TKA. Gait retraining has been shown to increase the number of modules and improve the quality of the modular organization and timing in individuals with post-stroke hemiparesis (52) and reduce co-contraction and knee pain in individuals with KOA (53). Therefore, rehabilitation protocols aimed at appropriately timed muscle activity may be necessary to restore independent activation of these muscle groups in individuals following a TKA.

In contrast to previous studies which have reported associations between modular organization and functional performance in impaired populations (23,35), the number of modules required to reconstruct the EMG of the participants with KOA was not associated with the clinical assessment measures, with the exception of a moderate correlation with their self-reported pain on the KOOS. However, a limitation of this pilot study is the small sample size. Though not statistically significant, there were trends of better average performance-based outcomes and KOOS symptom, ADL, and QOL subscales with a greater number of modules (Table 8). Thus, a larger study with a sufficient sample size in each module group is necessary to determine if differences in modular control complexity are associated with differences in measured or self-reported function in individuals with KOA. In addition, only EMG from the involved limb of the KOA group was included in this pilot study. While symmetry is a reasonable assumption in healthy adults, KOA commonly affects one knee more than the other. There may be differences in modular control between the involved limb and the contralateral limb which could provide insight into the relationship between clinical scores and motor control complexity.

In conclusion, this study revealed alterations in the modular control of walking in the presence of KOA. Though modular control complexity and organization was similar between healthy age groups, the participants with KOA exhibited decreased module complexity and altered module composition characterized by increased muscle co-activity, particularly of the quadriceps and hamstrings. This loss of modular control complexity may contribute to slower walking speeds, altered joint kinematics and kinetics, and increased self-reported pain. The findings of this pilot study suggest a change in underlying neural control strategy associated with the presence of KOA. Future work is needed to clarify the potential relationship between KOA-related changes in modular control strategy and functional outcomes.

## Data Availability

The data is not currently publicly available.

## ACKNOWLEDGEMENTS

The authors thank Dr. Jackie Lewis Devine and Dr. Greg Freisinger for their assistance with data collection.

